# Challenges in Tracking the Risk of COVID-19 in Bangladesh: Evaluation of A Novel Method

**DOI:** 10.1101/2021.08.03.21261567

**Authors:** Md. Enamul Hoque, Md. Shariful Islam, Arnab Sen Sharma, Rashedul Islam, Mohammad Ruhul Amin

## Abstract

Identifying actual risk zones in a country where the overall test positive rate (TPR) is higher than 5% is crucial to contain the pandemic. However, TPR-based risk zoning methods are debatable since they do not consider the rate of infection in an area and thus, it has been observed to overestimate the risk. Similarly, the rate of infection in an area has been noticed to underestimate the risk of COVID-19 spreading for the zones with higher TPR. In this article, we discuss the shortcomings of currently available risk zoning methods that are followed in the lower-middle-income countries (LMIC), especially in Bangladesh. We then propose to determine a risk zone by combining the rate of infection with TPR and effective reproduction number, *R*_*t*_ in a distinct manner from existing methods. We evaluate the efficacy of the proposed method with respect to the mass-movement events and show its application to track the evolution of COVID-19 pandemic by identifying the risk zones over time. Demo website for the visualization of the analysis can be found at: http://erdos.dsm.fordham.edu:3000

**CCS CONCEPTS:**

- **Applied computing** → **Health informatics**.

**ACM Reference Format:** Md. Enamul Hoque, Md. Shariful Islam, Arnab Sen Sharma, Rashedul Islam, and Mohammad Ruhul Amin. 2021. Challenges in Tracking the Risk of COVID-19 in Bangladesh: Evaluation of A Novel Method. In *Proceedings of August 15 (KDD Workshop on Data-driven Humanitarian Mapping, 27th ACM SIGKDD Conference)*. ACM, New York, NY, USA, 7 pages.

## 1 INTRODUCTION

The entire human race has been encountering a persistently mutating COVID-19 outbreak since December 2019. Although numerous studies reported the global domination of Wuhan-1 D614G lineage, new variants such as 501Y.V2 (B.1.351) received less attention, despite their severe transmission rate in many countries during the last winter season. This South African variant, 501Y.V2 (B.1.351), is able to reduce the neutralization sensitivity to convalescent plasma and therefore, it is capable of escaping from neutralization becoming more resistant to commercial SARS-CoV-2 vaccines [1]. On the other hand, the new Indian COVID-19 strain (B.1.617) called Delta variant, has caused India’s surge in COVID-19 infection cases and became a “variant of concern” as it has already spread over 40 nations, including the United Kingdom, Nepal and Singapore [2]. Notably, it has been observed that new COVID-19 variants are bringing a series of pandemic waves over time and therefore, identifying the COVID-19 risk zones in advance could play a pivotal role to contain the pandemic.

Unlike many other countries, Bangladesh has been braving the current pandemic by which more than 1.28 million people got infected and more than 21,162 people died as of August 3, 2021. Despite inadequate health care systems, shortage of physicians-nurses, and off-guard dwellers, Bangladesh seemed to contain the pandemic until February 2021 [3]. Unfortunately, we witnessed a surge in COVID-19 active cases from mid-February till the end of April 2021. Several factors might have contributed toward this surge: (1) The UK variant, South African variant and Indian variant of COVID-19 were found to spread the infectious disease at the community level [4, 5]; (2) reluctance to wear face-mask and maintaining social distancing were observed at the mass level [6]; (3) higher cost of COVID-19 test ($∼40) made it unreachable for the lower-income people having an income threshold of $1,025 per year [7]; and (4) first generation vaccines are rendered inefficient against Delta COVID-19 variants [1], among others.

It has been discussed among scientists all over the world and reasonably agreed upon that COVID-19 crisis might stretch over several years [8]. Hence, it is of utmost importance that we learn to live with it by adapting widely accepted public health recommendations at all levels. Communicating health advisory based on surveillance data and scientific evidence is the prerequisite to motivate the general population for wearing masks, maintaining social distance, taking vaccines, and overall living a healthy life during the pandemic. However, the lack of adequate COVID-19 tests and massive vaccination projects, and difficulty in tracing the asymptomatic carriers make it an uphill battle to contain the pandemic in Bangladesh like elsewhere. Furthermore, as the physical geography of the country differs across the largest delta, the native population of different zones have different lifestyles, making it difficult to implement an intervention plan.

Identifying potential risk zones is pivotal to contain the pandemic. However, TPR-based risk zoning methods are questionable since they do not account for the total number of infections in an area [9], and thus failing to reflect the actual scenario. In this paper, we propose to evaluate the risk of a zone by combining the daily cases per 100K population with TPR and effective reproduction number, *R*_*t*_. Since changes in TPR and *R*_*t*_ are very slow but cause rapid growths or decays in COVID-19 cases, we expressed their impact by a mathematical exponent. Based on the risk score, we clustered the districts of Bangladesh into four color-coded zones, such as red (tipping), orange (accelerated), yellow (community), and green (trivial). We discuss the following contributions in this paper:

- We study the evaluation of current risk zone indicator, and corresponding intervention plan in Bangladesh.
- We present a novel method of computing the risk zones, and show its efficacy compared to the existing methods.
- We discuss the association between the appearance of COVID-19 variants and their impacts on the spread of virus infection in different parts of the country.

## 2 RELATED WORKS

Multiple evidence suggested various insightful works that focused on the risk scoring of a zone for COVID-19 pandemic. For example, Arenas et al. [10] proposed a mathematical model that uses mobility flow and demography of the population to estimate the risk of municipalities in Brazil, whereas a network of researchers convened by Harvard’s Global Health Institute and the Edmond J. Safra Center for Ethics launched a framework, *Key Metrics for COVID-19 Suppression*, using daily cases per 100K people to label the risk of an area in one of 4 color codes (green, yellow, orange, red) [11]. Apart from these approaches, Chiu and colleagues used the geometric mean of TPR and reported cases of the previous 14 days to reliably estimate COVID-19 prevalence and transmission trends with a 7-day time lag across different states of the U.S.A[12].

Chande et al. measured the localized (county-level and state-level) risk associated with attending gatherings of different sizes in the United States with a simple binomial statistical model that takes 2 inputs (*p, n*), where *n* is the number of people attending the event and *p* is an estimation of per capita probability of COVID-19 infection [13]. Whereas, a more recent study called CovidActNow uses 3 metrics: daily new cases per 100K population, effective reproduction number (*R*_*t*_), and test positive rate (TPR) to assess the COVID-19 risk level across different states and counties in the United States [14].

In this paper, we present a novel method to compute the districtwise risk zoning and illustrate how the risk zoning helps to understand the progression of the third wave in Bangladesh in comparison to the previously identified risk zones of the past two waves. The proposed method differs from both the recently published methods of Chande et al. [13] and CovidActNow [14]. While Chande et al. measured the risk of attending an event by the number of people gathered in a particular area and CovidActNow computes the risk of an area based on hierarchical thresholding of three different metrics mentioned above, the proposed method results in a single risk score for any area by combining the same three metrics of CovidActNow. Our study shows that, in Bangladesh, the increase in the rate of infection in the third wave has started from border districts, evident by the increased identification of the *Delta* variant like different states in India; whereas past spikes started from the densely populated financial districts and due to international travelers.

## 3 METHODOLOGIES

### 3.1 Dataset

We collected the number of COVID-19 RT-PCR tests, positive cases, and death data of each district in Bangladesh on a daily basis from the Institute of Epidemiology Disease Control and Research (IEDCR) website for the analysis presented in this paper. In addition to this, we collected COVID-19 status reports for each country from the Our World In Data (OWID) website^1^. Also, SARS-CoV-2 sequences submitted from Bangladesh till May 2021 were obtained from GISAID (PMCID: PMC5388101). We used *NextClade*^2^ to call the mutations and clades against the MN908947 reference (PMCID: PMC6247931).

### 3.2 A Novel Risk Assessment Method

To evaluate the risk of COVID-19 pandemic, World Health Organization (WHO) and European Commission (EC) have suggested to use the formalism shown below [15]. The Brown School of Public Health (BSPH) further extended the proposal with a cutoff for the 4 color-coded risk zones as per the guideline of *Key Metrics for COVID Suppression* [11].

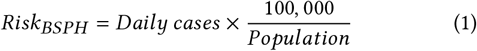

The test positivity rate (TPR) based risk zone definition is shown below [16]. The Bangladesh government uses a TPR-based risk zoning in policy making.

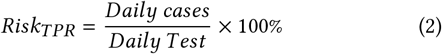

We have observed that the TPR is not sufficiently low (< 5%) in most of the global south countries. This gives rise to the question whether BSPH risk evaluation will be sufficient if the TPR is high and vice versa. To assess this question, we analyzed BSPH, TPR, and R_t_ for the reported COVID-19 testing rate of countries categorized into 5 INFORM classes^3^. If a country has a higher INFORM score then it means it has a lower capacity to manage humanitarian crisis or disaster situations. To show how the risk zoning is impacted by the the BSPH, *R*_*t*_, and TPR, we present a set of scatter plots in the **Figure 1**. These plots represent that countries of higher INFORM risk classes are doing much better in managing the COVID-19 pandemic than that of lower INFORM risk class countries, meaning BSPH is underestimating the risk. On the other hand, if we consider TPR-based risk, then we observe that countries in the medium INFORM risk class fall into higher risk zones when compared with the lower INFORM risk class countries that fall into a low-risk zone due to their testing rate. In later section, we will show how TPR is overestimating the risk zoning at a district level for Bangladesh.

**Figure 1:**
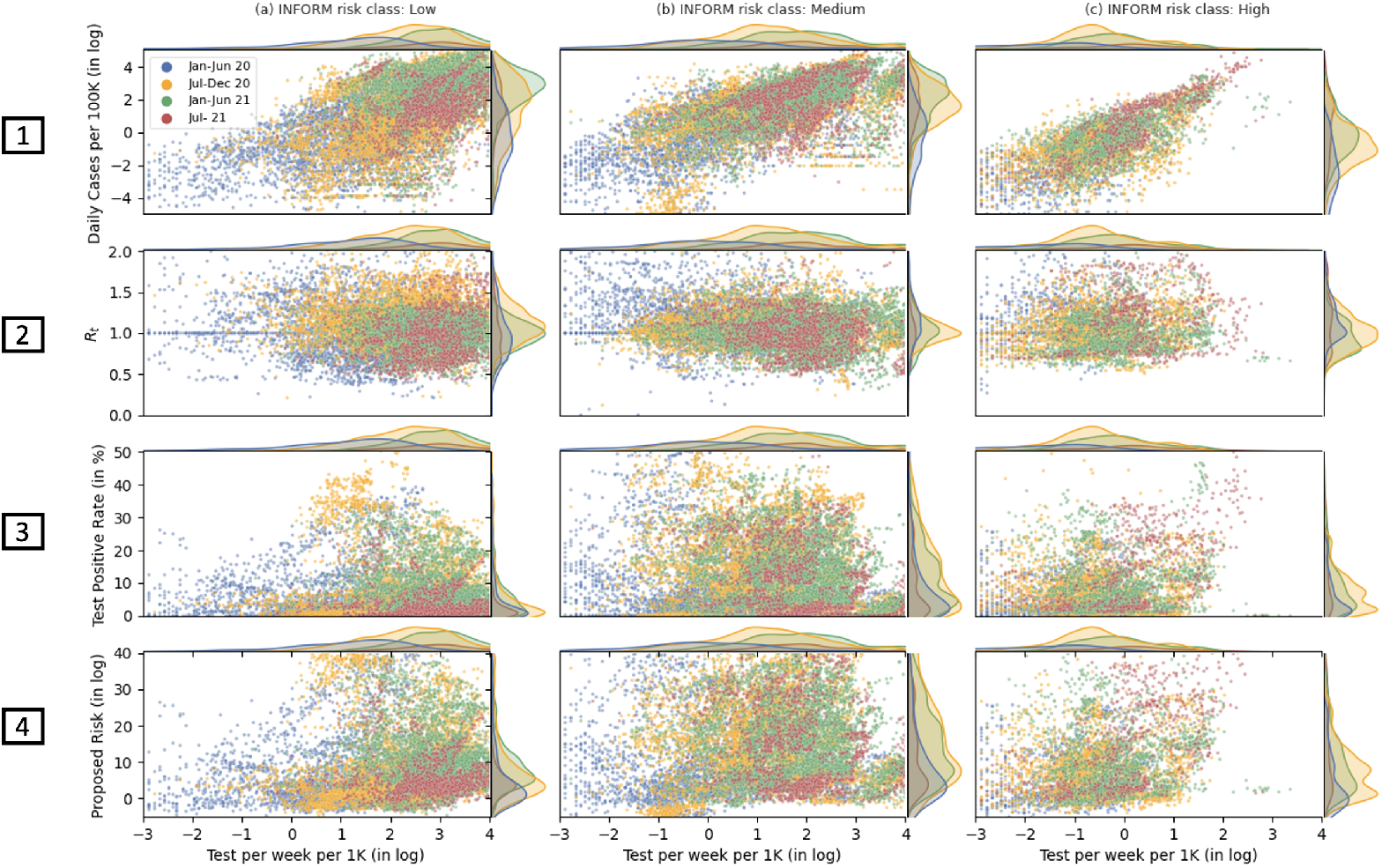
Scatter plots presenting BSPH (daily cases per 100K), *R*_*t*_, TPR, and proposed risk score with respect to testing rate as reported in the OWID website for countries categorized into 3 of the 5 INFORM classes. Two extremes of these 5 INFORM classes, i.e., *very low* and *very high*, were excluded in this plot due to the lack of data. The plot shows that higher countries with a higher INFORM risk reported lower number of daily COVID-19 cases and higher number of TPR. So, according to BSPH these countries fall into low risk zone, but according to the TPR the assessment reach to a contradictory conclusion. Whereas, *R*_*t*_ does not vary a lot for any of the countries. On the other hand, our proposed risk score shows that higher INFORM class countries, in fact, have higher risk scores due to the lower testing rates.

In our approach of the risk evaluation, we defined the *localized severity* by considering both the BSPH and an exponent of the TPR. Here, note that as TPR ranges between 0 to 1 and often close to 0, the exponent of TPR is equivalent to (1 + *TPR*). Thus for lower INFORM risk class countries where TPR is close to 0, *localized severity* remains almost similar to BSPH, but it changes to higher value for the low INFORM countries where the TPR is higher. Consequently we define the severity as,

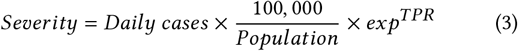

Since the transmission risk relies on the number of available infection in a community, we need to approximate the available but not tested positive cases. Finding the probable cases, we depend on the effective reproduction number *R*_*t*_ of that location as it provides an estimation of the number of probable cases in the next time interval. Hence, to approximate the immediate risk, we define the sensitivity as,

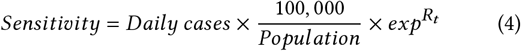

Due to the incubation period (or the serial interval in *R*_*t*_), the number of available infection in a community should be much higher than that directly provided by the *R*_*t*_. With above consideration, we define the risk of an area as,

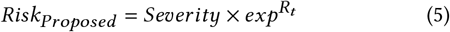

Thus, the proposed risk formalism includes the tested positive cases as well as the probable cases that are not tested and will be detected in the next interval for a community in an outbreak. The fourth horizontal panel in the **Figure 1** shows the outcome of the proposed risk scoring method. It clearly shows that the higher INFORM class countries actually fall into higher risk zone.

In epidemic, the growth or decline are often described as the exponential rate of change [17]. As the test positivity rate (TPR) and the effective reproduction number (*R*_*t*_) are the spontaneous terms which may surge or decline very fast with time, we integrated the exponential term with these parameters. We present the detail of risk zone method and corresponding score cut-offs for defining a color code in **Table 1**.

**Table 1:** The 4 color coded COVID-19 risk zone definition.

| Zone Code | Green | Yellow | Orange | Red |
| --- | --- | --- | --- | --- |
| Zone Definition | Trivial | Community Spread | Accelerated Spread | Tipping |
| Meaning of Color Code | Close to Containment | Potential Spreading | Escalating Spreading | Unchecked Spreading |
| Zoning by BSPH | Less than 1 | 1 to less than 9 | 9 to less than 24 | 24 and more |
| Zoning by TPR | Less than 10% | 10% to less than 20% | 20% to less than 30% | 30% and more |
| Zoning by Proposed | Less than 1 | 1 to less than 9 | 9 to less than 24 | 24 and more |

## 4 RISK SCORING METHODS EVALUATION

In the heatmap of **Figure 2**, we have shown the dynamics of the proposed risk method along with the standard methods based on cases, TPR, Rt and CovidActNow for the last year of COVID-19 pandemic in Bangladesh. The districts are arranged from the slightly industrialized district Netrakona to the highly industrialized capital district Dhaka to observe the impact of the COVID-19. It needs to be mentioned that the testing rate (the number of test per week per 1,000 population) is less than 1 for all the districts with mostly less than 0.001. We used four different risk zones, namely trivial (green), community spread (yellow), accelerated spread (orange) and tipping point (red) for presenting corresponding risk of a district. The heat map in **Figure 2** demonstrates that TPR-based method is overestimating the risk score of a zone relative to all other methods. For example, Netrakona, Sherpur, Kurigram district hardly showed any risk throughout the entire 2020 according to BSPH, *R*_*t*_, CovidActNow and the proposed method, whereas TPR-based method classifies those districts as high risk zones for COVID-19 transmission multiple times through the past year. On the other hand, BSPH-based risk zoning is underestimating the risk of COVID-19 infection in Dhaka city, one of the most densely populated cities in the world. Dhaka is considered as the epicenter of COVID-19 diffusion in Bangladesh as it has been reported in numerous papers [18]. BSPH method has also resulted in lower risk score for several other cities including Chittagong, Narayanganj, and Gazipur.

**Figure 2:**
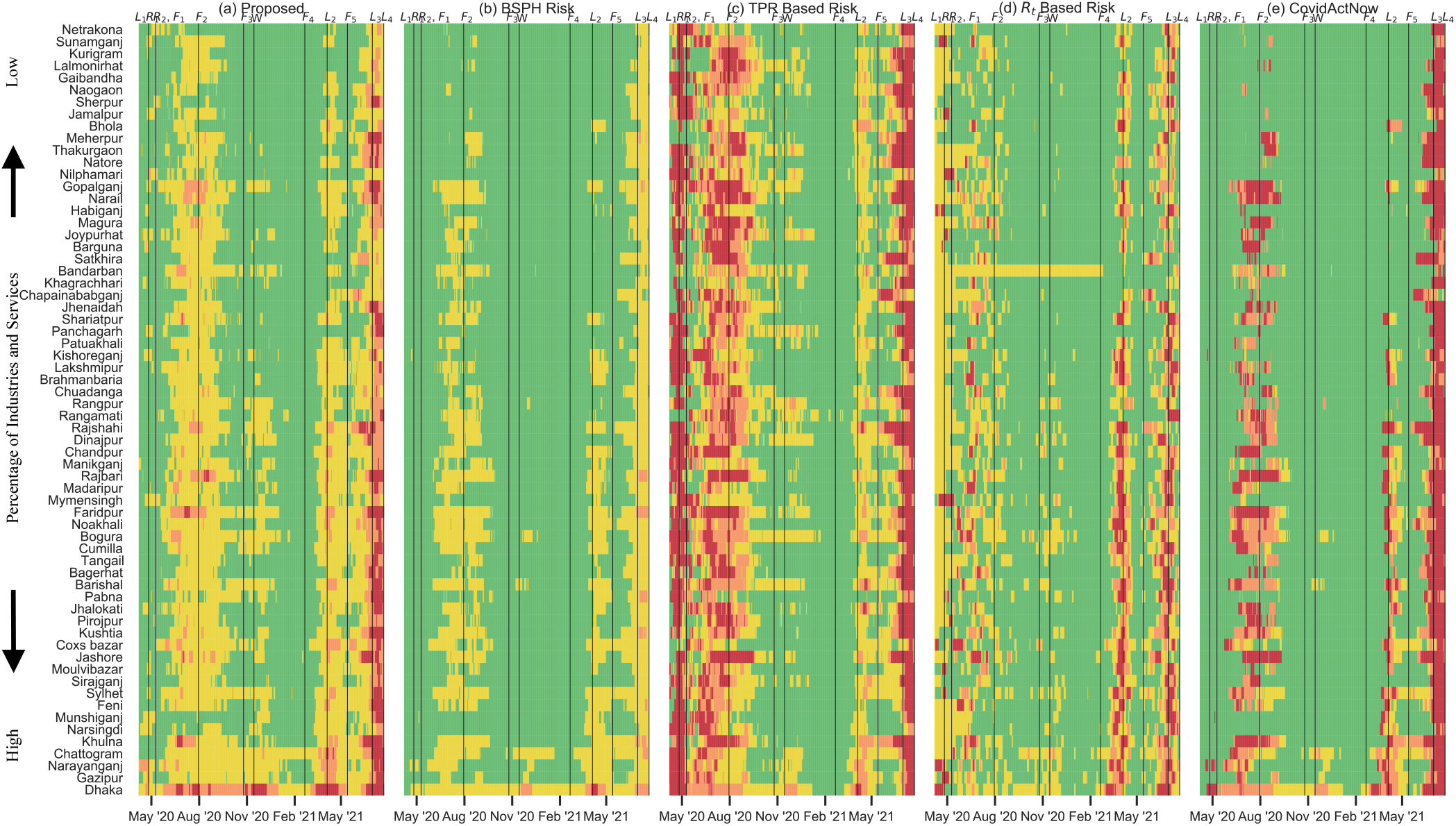
The dynamics of the 4 color coded risk zone for COVID-19 evaluated by (a) Proposed, (b) BSPH, (c) TPR based, (d) *R*_*t*_ based, and (e) CovidActNow methods. The vertical lines represents the major events during the pandemic where mass movement took place before and after each lockdown (*L*_*1*_ *–L*_*2*_) or due to public festivals (*F*_*1*_ *–F*_*5*_)

The risk dynamics using *R*_*t*,_ is considered to be a good approximation to the infectiousness overtime. Unfortunately, due to the very low testing rate in Bangladesh, we have observed comparatively lower number of detected cases for which *R*_*t*_ resulted in very different outcome than that of BSPH or any other method. Besides, the K-sys formalism of the computation of *R*_*t*_ provides less confidence in lower number of detected cases [18]. Whereas, CovidActNow provides a risk metric based on hierarchical thresholding of daily cases per 100K (same as BSPH), TPR, and *R*_*t*_, i.e. if daily cases per 100K in a zone is within green range, then risk of that zone is green, otherwise the maximum risk score among daily cases per 100K, TPR, and *R*_*t*_ is considered to define the zone color code. Now, since BSPH method has a tendency to underestimate the risk in some places and CovidActNow simply ignores other metrics in case the BSPH indicates a zone risk as green, CovidActNow has a tendency to underestimate the risk of a zone in some cases. Moreover, in **Figure 2**, we observe that the rise and decline in the number of districts in the red zone according to CovidActNow highly correlate with that of BSPH. But unfortunately, for each yellow zone as defined in BSPH, CovidActNow computed a very high scores due to higher corresponding TPR; and thus, it should be considered as a weakness of this method. In other words, as CovidActNow depends on hierarchical thresholding of other three metric discussed above, in the process, it primarily inherits the overestimation characteristics of TPR. In our proposed method, the risk scoring method combined BSPH, TPR and *R*_*t*_ all together to capture the severity of COVID-19 infection as well as to make it sensitive for immediate future risk. In this distinct approach, the risk score of each district correlate with that of BSPH method but it ensures to avoid both the overestimation and underestimation phenomena we discussed above. Moreover, by considering the risk zone cut-offs from the BSPH formalism, we obtained a more appropriate risk zone for the districts in terms of the major events.

We present the evolution of the risk zones among the 64 districts of Bangladesh in **Figure 3**. We integrated the major events when mass movement took place within the country at different times of the year. We observe that TPR has overestimated the risk zones at the very beginning even before the first lockdown was declared, and then during the lockdown but failed to capture the second lockdown. Similar to TPR, CovidActNow overestimated risk zones due to its hierarchical decision process. On the other hand, BSPH underestimated the risk of districts and only elevated risk to yellow for some districts during first and second lockdown. It failed to capture the severity of pandemic in almost all the densely populated financial districts. Besides, *R*_*t*_ demonstrative its sensitivity only towards financial districts by elevating corresponding risk zones while failing to determine the appropriate risk zones of other districts, such as yellow zones. The proposed method on the other hand captured the severity of pandemic and could relate with the major mass movement events, such as first and second lockdown (*L*), restarting the economy (*R*), and religious festivals (*F*).

**Figure 3:**
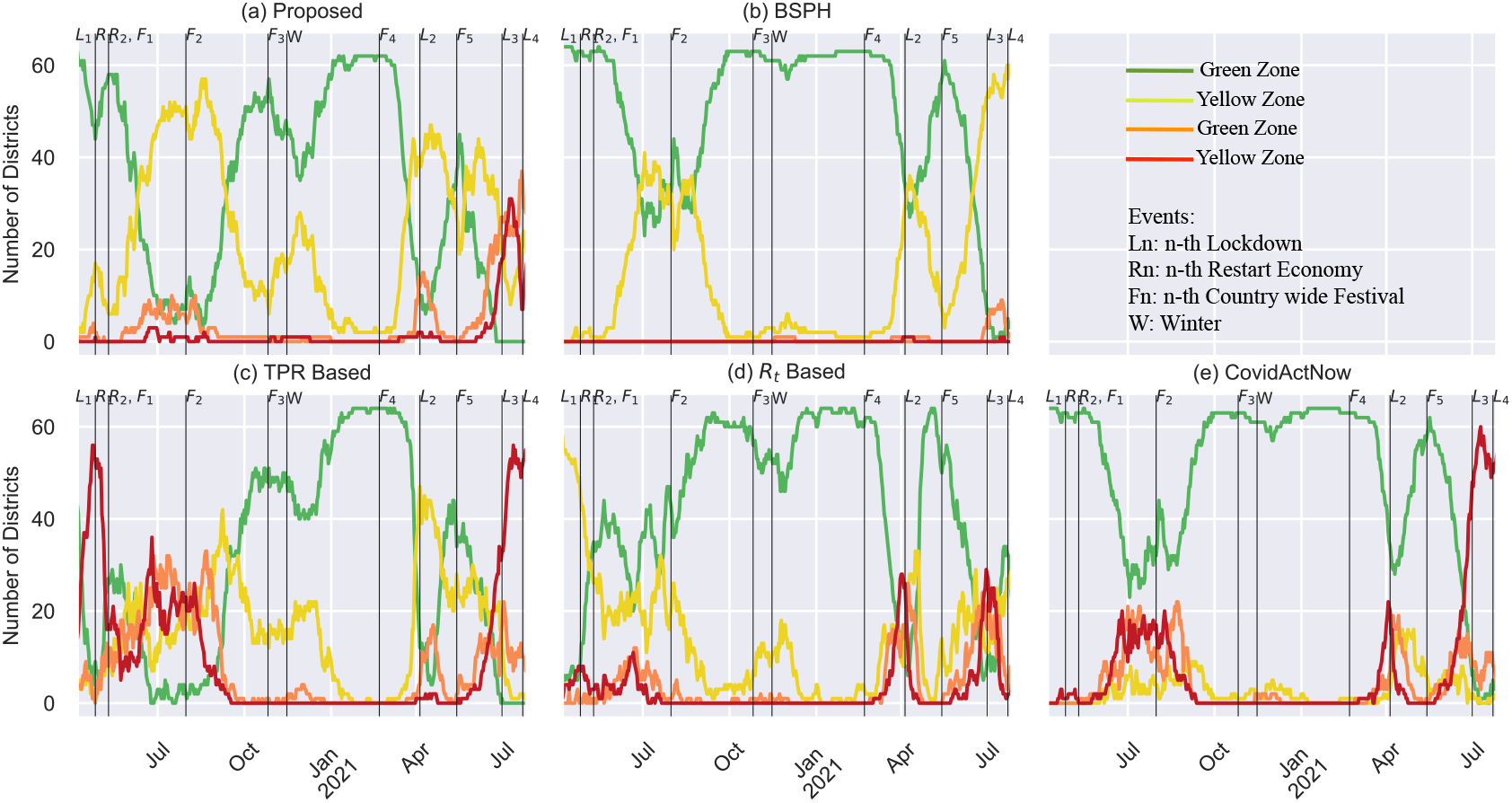
The evolution of the risk zones as evaluated by (a) Proposed, (b) BSPH, (c) TPR based, (d) *R*_*t*_ based, and (e) CovidActNow methods. The major events that contributes to mass movement in Bangladesh are indicated using vertical lines. We choice to compare the lockdown *L*, restarting economy *R*, country-wide festival *F* and the winter *W*.

Finally, we present the district-wise risk zoning and present the comparison of current situation in **Figure 4(C)** with previously identified risk zones in the past lockdown as shown in **Figure 4(a) and (b)**. It shows that latest increase in the rate of infection started from border districts of Bangladesh due to the surge in COVID-19 cases in India; whereas past incidents started from the financial district Dhaka due to international travelers.

**Figure 4:**
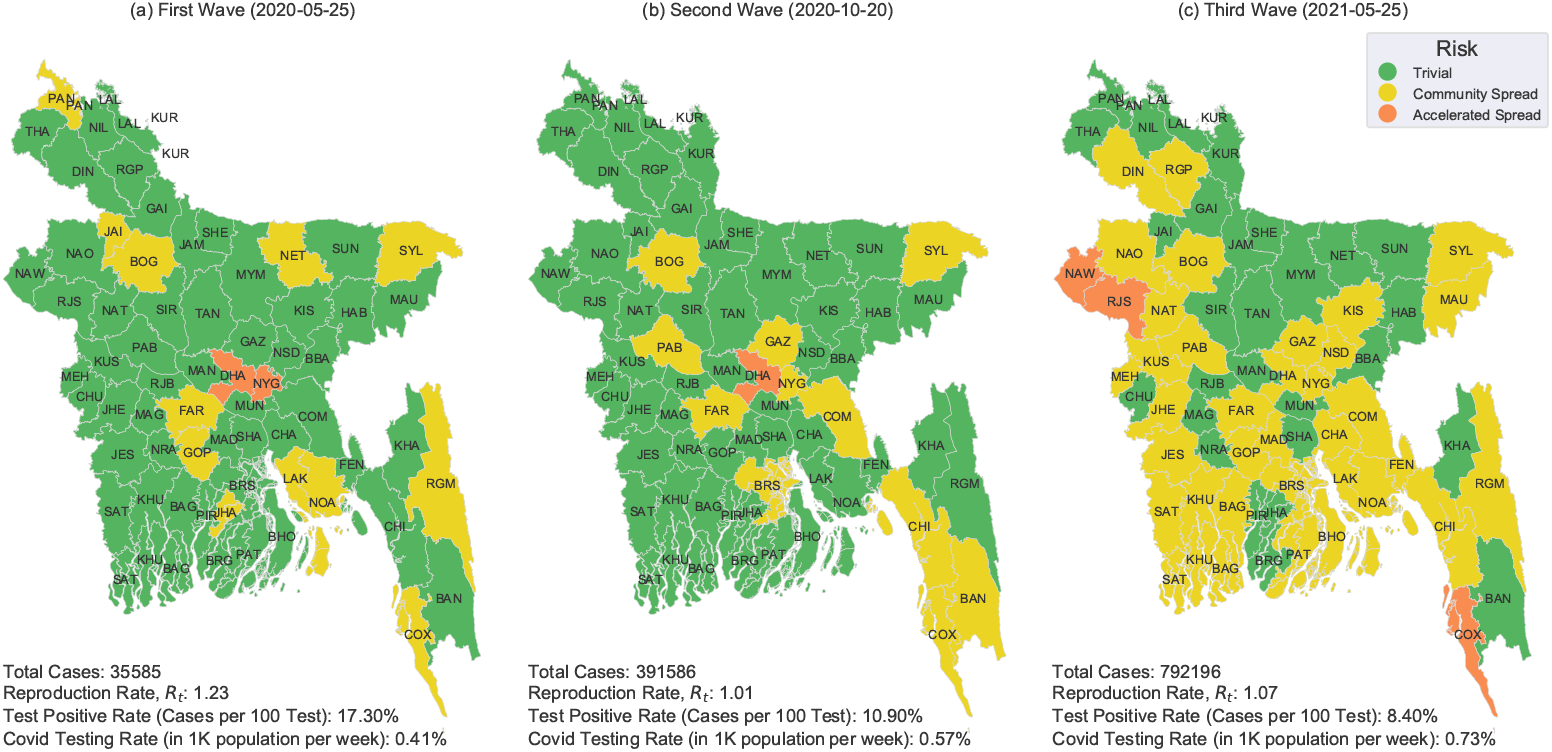
The district-wise risk zoning of Bangladesh shows that the primary high infection occurs at Dhaka for the (a) first and (b) second waves. But there is high infection rate at the Bangladesh-India border for the (c) third wave.

## 5 EMERGING COVID-19 VARIANTS ARE ASSOCIATED WITH RISK

SARS-CoV-2 sequences submitted from Bangladesh till May 2021 were obtained from GISAID (PMCID: PMC5388101). We analyzed these sequences between April 2020 to May 2021 and associated variant distributions with risk. In the year of 2020, two variant clades e.g., 20A and 20B were dominating in Bangladesh as shown in **Figure 5**. Since the beginning of 2021, the South African variant 20H/501Y.V2 (B.1.351) continued to dominate the country. Notably, the first and second wave of the pandemic were associated with the emergence of variant clade 20A and 20H/501Y.V2 (B.1.351) respectively [19]. However, recent emergence of the more contagious Delta variant (B.1.617.2) in India influenced the variant distribution in June 2021 where 21A (B.1.617.2) clade has become the most dominant type (46% of the sequences) in Bangladesh. In June 2021, the most frequent variant observed in Bangladesh was B.1.617.2, which had been reported earlier to be dominant in Maharashtra and West Bengal of India [20, 21]. Whereas, a month before that, in May 2021, the most frequent variant in Bangladesh was B.1.351.3, a new strain originally identified in South Africa (see **Table 2**) [19]. Recently, Chapainawabgonj (marked as ‘NAW’ in **Figure 4(c)**), a western district of Bangladesh that shares the border with West Bengal of India, identified many COVID-19 cases where 88% (7/9) sequences were identified as B.1.617.2 (Delta variant). Since Chapainawabgonj does not have any international airport, the B.1.617.2 variant might have been introduced in Bangladesh through road communications across the border (see **Figure 4(c)**). Thus, based on day-to-day data, our proposed method has the potential to track unanticipated spread of COVID-19 cases potentially caused by a transmissible variant at district level.

**Figure 5:**
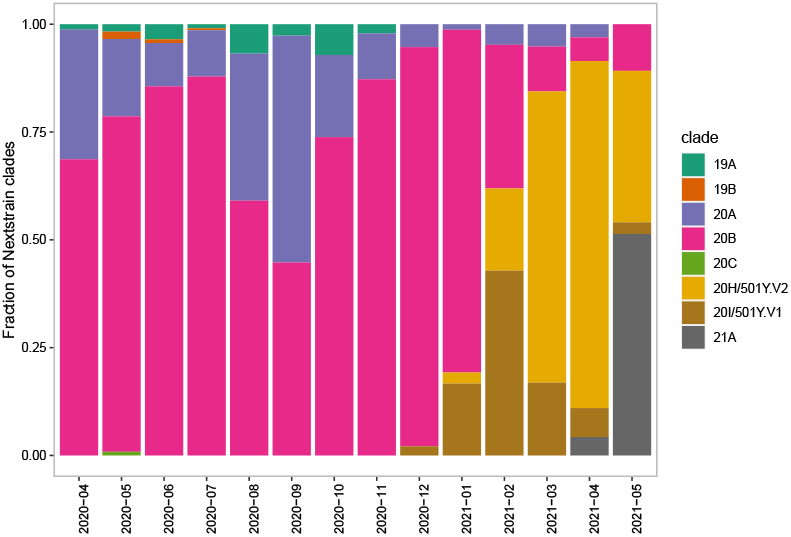
Fraction of different COVID-19 variants that appeared throughout the year 2020-2021 in Bangladesh.

**Table 2:** Five most frequent variant groups in May and June 2021 in Bangladesh.

| Pangolin_lineage | Count | Frequency | Month, 2021 |
| --- | --- | --- | --- |
| **B.1.351.3** | **118** | **0.72** | **May** |
| B.1.351 | 18 | 0.11 |  |
| B.1.1.7 | 10 | 0.061 |  |
| B.1.1.318 | 8 | 0.0488 |  |
| B.1.525 | 5 | 0.0305 |  |
| **B.1.617.2** | **17** | **0.459** | **June** |
| B.1.351.3 | 18 | 0.11 |  |
| B.1.1.318 | 3 | 0.0811 |  |
| B.1 | 2 | 0.0541 |  |
| B.1.1.220 | 1 | 0.027 |  |

## 6 CONCLUSION

The proposed method for zoning will provide alternative measures for making policies on zonal lockdown for COVID-19 crisis management. Our analysis shows that Bangladesh is on the verge of a third wave of COVID-19 infection due to the Indian variant of the COVID-19 virus. One may assume that this might be the beginning of a new humanitarian crisis in Bangladesh where the poverty increased by 20% in last one year [22]. Hence, increasing COVID-19 test at the population level, and collecting the COVID-19 patient data on a daily basis without any delay from all over the country should be the first couple of steps to combat the current humanitarian emergency. This will facilitate making a reliable forecast about the progression of the outbreak and thus, in designing proper control methods. Furthermore, we propose to implement this zoning method at a higher resolution geographical boundary, i.e, thana, union, or ward level, to revive the economy of the country while saving hundreds of thousands of lives.

## Data Availability

We collected the number of COVID-19 RT-PCR tests, positive cases, and death data of each district in Bangladesh on a daily basis from the Institute of Epidemiology Disease Control and Research (IEDCR) website for the analysis presented in this paper. In addition to this, we collected COVID-19 status reports for each country from the Our World In Data (OWID) website. Also, SARS-CoV-2 sequences submitted from Bangladesh till May 2021 were obtained from GISAID (PMCID: PMC5388101).

## ACKNOWLEDGMENTS

We sincerely thank to the Access to Innovation (a2i) Programme, Institute of Epidemiology Disease Control and Research, and Ministry of Health and Welfare of the Government of Bangladesh to provide the necessary data to perform this study.

## Footnotes

1 https://ourworldindata.org

2 https://clades.nextstrain.org/

3 https://drmkc.jrc.ec.europa.eu/inform-index

## REFERENCES

[1] Pengfei Wang, Ryan G Casner, Manoj S Nair, Maple Wang, Jian Yu, Gabriele Cerutti, Lihong Liu, Peter D Kwong, Yaoxing Huang, Lawrence Shapiro, et al. Increased resistance of sars-cov-2 variant p. 1 to antibody neutralization. Cell host & microbe, 29(5):747–751, 2021.

[2] Gayathri Vaidyanathan. Coronavirus variants are spreading in india—what scientists know so far. Nature, 593(7859):321–322, 2021.

[3] Zakaria Shams Siam, Md Arifuzzaman, Md Salik Ahmed, Faisal Ahamed Khan, Md Harunur Rashid, and Md Shariful Islam. Dynamics of covid-19 transmission in dhaka and chittagong: Two business hubs of bangladesh. Clinical epidemiology and global health, 10:100684, 2021.

[4] Ovinu Kibria Islam, Hassan M Al-Emran, Md Shazid Hasan, Azraf Anwar, Md Iqbal Kabir Jahid, and Md Anwar Hossain. Emergence of european and north american mutant variants of sars-cov-2 in south-east asia. Transboundary and emerging diseases, 68(2):824–832, 2021.

[5] Amena Ahmed Moona, Sohel Daria, Md Asaduzzaman, and Md Rabiul Islam. Bangladesh reported delta variant of coronavirus among its citizen: Actionable items to tackle the potential massive third wave. Infection Prevention in Practice, 2021.

[6] Md Taimur Islam, Anup Kumar Talukder, Md Nurealam Siddiqui, and Tofazzal Islam. Tackling the covid-19 pandemic: The bangladesh perspective. Journal of public health research, 9(4), 2020.

[7] Sophie Cousins. Bangladesh’s covid-19 testing criticised. The Lancet, 396(10251):591, 2020.

[8] OECD. Beyond containment: Health systems responses to covid-19 in the oecd. 2020.

[9] Ensheng Dong, Hongru Du, and Lauren Gardner. An interactive web-based dashboard to track covid-19 in real time. The Lancet infectious diseases, 20(5):533–534, 2020.

[10] Alex Arenas, Wesley Cota, Jesús Gómez-Gardenes, Sergio Gómez, Clara Granell, Joan T Matamalas, David Soriano-Panos, and Benjamin Steinegger. A mathematical model for the spatiotemporal epidemic spreading of covid19. MedRxiv, 2020.

[11] Key Metrics for COVID Suppression, last accessed: June 3.

[12] Martial L Ndeffo-Mbah et al. Using test positivity and reported case rates to estimate state-level covid-19 prevalence in the united states. medRxiv, 2020.

[13] Aroon Chande, Seolha Lee, Mallory Harris, Quan Nguyen, Stephen J Beckett, Troy Hilley, Clio Andris, and Joshua S Weitz. Real-time, interactive website for us-county-level covid-19 event risk assessment. Nature Human Behaviour, 4(12):1313–1319, 2020.

[14] U.S. COVID Risk & Vaccine Tracker, last accessed: June 3, 2021.

[15] INFORM Covid-19 Warning Datasources, last accessed: June 3, 2021.

[16] COVID Data Tracker, last accessed: June 3, 2021.

[17] Junling Ma. Estimating epidemic exponential growth rate and basic reproduction number. Infectious Disease Modelling, 5:129–141, 2020.

[18] Md Shariful Islam, Md Enamul Hoque, and Mohammad Ruhul Amin. Integration of kalman filter in the epidemiological model: a robust approach to predict covid-19 outbreak in bangladesh. International Journal of Modern Physics C, page 2150108, 2021.

[19] Senjuti Saha, Arif M Tanmoy, Yogesh Hooda, Afroza Akter Tanni, Sharmistha Goswami, Syed Muktadir Al Sium, Mohammad Saiful Islam Sajib, Roly Malaker, Shuborno Islam, Hafizur Rahman, et al. Covid-19 rise in bangladesh correlates with increasing detection of b. 1.351 variant, 2021.

[20] Sarah Cherian, Varsha Potdar, Santosh Jadhav, Pragya Yadav, Nivedita Gupta, Mousmi Das, Partha Rakshit, Sujeet Singh, Priya Abraham, Samiran Panda, et al. Convergent evolution of sars-cov-2 spike mutations, l452r, e484q and p681r, in the second wave of covid-19 in maharashtra, india. bioRxiv, 2021.

[21] Houriiyah Tegally, Eduan Wilkinson, Marta Giovanetti, Arash Iranzadeh, Vagner Fonseca, Jennifer Giandhari, Deelan Doolabh, Sureshnee Pillay, Emmanuel James San, Nokukhanya Msomi, et al. Detection of a sars-cov-2 variant of concern in south africa. Nature, 592(7854):438–443, 2021.

[22] Survey: New poor in Bangladesh stands at 24.5 million, last accessed: April 20, 2021.

